# Predictors and outcome of post-stroke depression among adults admitted for first stroke at referral hospitals in Dodoma, Tanzania: a protocol for a prospective longitudinal observational study

**DOI:** 10.1101/2025.01.05.25320012

**Authors:** Sadiki Mandari, Azan Nyundo

**Author notes:** Corresponding author Azan Nyundo P O Box 395 Dodoma 1 TIBA STREET, 41218 IYUMBU, DODOMA TANZANIA. **Funding statement:** This project did not receive any funding. **Data availability statement:** Data will be available and shared as per agreement of terms and conditions once the data collection is completed. **Competing interest statement:** The authors declare there is no conflict of interest.

## Abstract

**Background:** Survivors of strokes are prone to disabilities, especially in underdeveloped countries. Post-stroke depression (PSD) is a common neuropsychiatric condition that exacerbates symptoms and raises the danger of stroke recurrence, disability, and mortality. Nevertheless, little is documented about PSD’s incidence, predictors, and consequences.

This study aims to assess predictors and outcomes of post-stroke depression among patients admitted with the first stroke episode at referral hospitals in Dodoma, Tanzania.

**Methods and analysis:** The study is a prospective longitudinal observational design; a consecutive sampling technique will be used to attain the estimated sample size. Adults aged ≥18 years who have had their first stroke episode, within 14 days, and the stroke diagnosis will be verified through brain imaging using CT or MRI. The study will be conducted at referral hospitals in Dodoma region, Tanzania. At admission, baseline clinical parameters will be recorded, and PSD will be evaluated at one and three months after a stroke. Data will be summarised using descriptive statistics; continuous data will be reported as mean (SD) or median (IQR) while categorical data as frequencies and proportions. The PSD predictors will be determined using logistic regression analysis. The study will adhere to data-sharing guidelines and take ethical considerations into account.

**Ethics and dissemination:** The University of Dodoma’s institutional Research Review and Ethical Committee has granted permission to conduct the study with reference number MA.84/261/02. The relevant authorities granted approval for the study to be carried out at DRRH and BMH.

## Introduction

Stroke is the second major cause of death globally [1] and is also linked to the development of psychiatric symptoms and impaired quality of life[2]. Similar to the developed world where majority of stroke survivors suffer form Ischaemic stroke, [1] approximately 80% of stroke in the develong countries is also Ischaemic [3]. The prevalence is the highest in Sub-Saharan Africa and other low and middle-income countries, including Tanzania; the incidence increased by 70% from 1990 to 2019, and the prevalence increased by 85%[4]. Despite the implementation of preventive measures and risk factor reduction similar to high-income countries, high rate of strokes is recorded in both urban and rural areas of Tanzania[5].

Following a stroke, there is a high risk of developing neuropsychiatric manifestations with up to a ten-fold increase in mortality rates [6]. A 20 to 50% one-month incidence of post-stroke depression is reported and persists for three to six months after the stroke [7]. While in high-income countries, the prevalence of PSD ranges from 25% to 79% [8], the prevalence rates of 32%, 54%, and 89% are observed in low- and middle-income countries of Uganda, the Democratic Republic of the Congo, and the Central African Republic, respectively.

Although there is a mixed evidence, several factors including female gender, advanced age, medical and psychiatric history, social support, and stroke-related factors, including severity and degree of disability have been accounted for PSD. [9].

The study aims to determine the prevalence, predictors and outcome of PSD among patients admitted with the first stroke episode at referral hospitals in Dodoma, Tanzania.

## Study aims

### Aim 1

To determine the baseline prevalence of post-stroke depression at one month among patients admitted with the first stroke episode at referral Hospitals in Dodoma.

### Aim 2

To determine predictors of post-stroke depression among patients admitted with the first stroke episode at referral Hospitals in Dodoma.

### Aim 3

To determine the outcome of post-stroke depressive symptoms at three months following the first stroke episode among patients admitted at referral Hospitals in Dodoma.

### Aim 4

To determine predictors of significant improvement of depressive symptoms among patients admitted with the first episode of a stroke at referral Hospitals in Dodoma.

## Methods and analysis

### Study design

The study will be a prospective longitudinal observation design.

### Study setting

The study will be conducted in Dodoma’s referral hospitals, the Dodoma Regional Referral Hospital (DRRH) and Benjamin Mkapa Hospital (BMH). Dodoma is Tanzania’s capital, with a population of 3,085,625 people per the 2022 national census [10]. The coverage includes referrals from all Dodoma districts of Mpwapwa, Bahi, Kongwa, Chemba, Kondoa and neighbouring regions. BMH has 400 beds while DRRH has 480 bed capacities, where all patients will be received at emergency department before being transferred to the respective unit or ward. Stroke is among the top listed conditions admitted at the the hospitals, with about 20 cases admitted every month in the medical ward and intensive care unit for those requiring close observation. Also, both BMH and DRRH has advanced radiological investigations, including CT- scan covering patients from neighbouring regions.

### Study population

Participants are adults aged 18 years or older admitted to the Internal Medicine wards of either DRRH or BMH with a diagnosis of the first stroke as per the World Health Organization, defined as “rapid development of clinical signs of focal or global disturbance of cerebral function lasting more than 24 hours or leading to death, with no apparent cause other than vascular origin” and confirmed by CT-scan or MRI, and the duration of symptoms lasting not more than 14 days[11].

### Inclusion criteria

- Patients aged 18 years or older admitted with the first episode of stroke.
- Capacity to provide informed consent or proxy consent from a close relative or custodian in case the patient is incapable.
- Patients with the first stroke episode within 14 days confirmed by CT-scan/MRI.

### Exclusion criteria

- All patients with severe sensory impairment (deafness and blindness) that will compromise the assessment of key dependent and independent variables.
- Patients with traumatic intracerebral haemorrhage.
- Patients with intracerebral haemorrhage due to tumour.
- Patients with transient ischemic attack (TIA).
- Patients with traumatic subarachnoid haemorrhage. Patients with a known history of chronic neurological disorders with an established risk of psychiatric manifestations such as epilepsy, multiple sclerosis, and neurogenerative disorders.

### Sample size calculation

The sample size will be estimated utilising formula for proportion in a prospective cohort study [12]

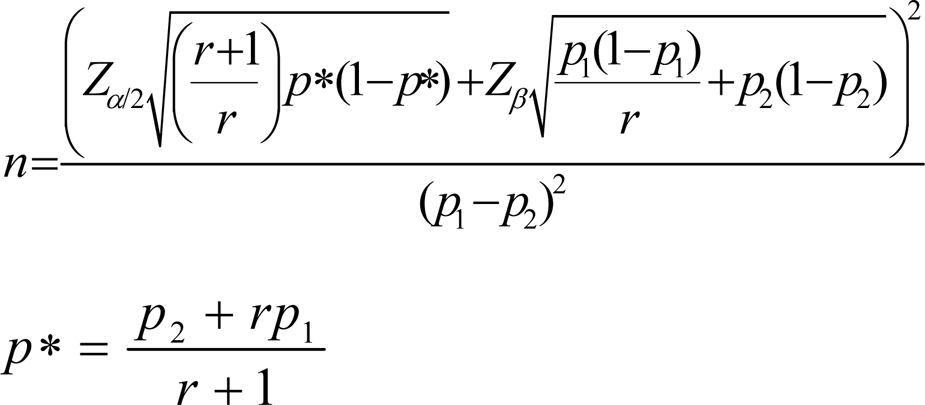

**Where by**

r = ratio between the two groups

p_1_ = PSD one-month prevalence (obtained from literature)

p_2_ = PSD one-moth prevalence observed or expected from the study

p_1_ – p_2_ = effect size

Z_β_ = standard normal variate for statistical power

Z_α/2_ = standard normal variate for significance level

The prevalence of PSD at 1 months is 50% [8]

The one-month prevalence in this study is expected to be 30%

Therefore;

r = 1.67

p_1_ = 50%

p_2_ = 30%

p_1_ – p_2_ = 20%

Z_β_ = 1.28 for statistical power of 90%

Z_α/2_ = 1.96 for significance level of 95%

Considering the 30 % attrition rate [13]

Therefore, the minimum sample size estimated is 274 patients.

### Sampling methods/technique

A consecutive sampling method will be used whereby the sample will be attained by selecting every available candidate meeting the inclusion criteria and admitted through the emergency/outpatient department to the wards until the desired sample size is reached.

### Data collection procedure/recruitment of patients

Direct interviews with the patient and/or immediate guardian will be used to collect personal information (including sex, age, marital, and occupation), past medical history (such as hypertension and diabetes mellitus), and lifestyle (alcohol drinking and smoking), whereby current smoking/alcohol use, defined as those who smoke or take alcohol within the last 12 months. Thorough history taking on symptoms and physical examination assessing the atrial fibrillation using ECG, Leukoaraiosis, stroke characteristics like stroke laterality, site of lesion using CT/MRI scan and stroke severity will be evaluated using the National Institute of Health Stroke Scale with a total score of 42-points will be computed to categorise stroke severity. The severity will be classified as mild stroke if the score is 1 to 4, 5 to 15 as moderate to severe stroke, 16 to 20 as severe stroke and 21 to 42 as very severe stroke [14]. Patients who will score ≤5 usually indicate a strong possibility for a good recovery with a sensitivity of 72% and specificity of 89% [15].

### Clinical examination

Blood pressure (BP) readings will be taken using an automated digital machine AD Medical Inc. brand; patient will be in a supine position with the arm placed at the same position as the heart; a minimum of two readings will be taken 2 minutes apart. The affected arm will be avoided in order to reduce false results. Hypertension will be defined as BP ≥140/90 mmHg in patients with a history of hypertension or on antihypertensive medications [16].

The radial pulse will be measured using a finger pulse oximeter model FL – 100, preferably on the limb unaffected for 1 minute. At least two medical doctors will confirm the presence of arrhythmia [17].

### Laboratory investigations

This study’s testing will be done in accordance with accepted DRRH and BMH standard operating procedures. A laboratory expert will request each participant’s consent to the venepuncture and finger prick before taking blood samples. Before sample collection tubes [EDTA (K2/K3) sodium fluoride plain, no modifications] will be labelled with the hospital registration number corresponding to the patient, and a tourniquet is applied proximally to the upper arm, which is about 6 cm above the elbow joint. Cleaning the region with 70% methylated spirit in a circular motion and letting it dry for 20 seconds before performing the venipuncture. A 10cc syringe will next be used to draw 5ml of venous blood from each arm for lipid analysis (high-density lipoprotein (HDL), low-density lipoprotein (LDL), total cholesterol, and triglycerides). The blood samples will be transported into a cool box to a recognised Dodoma regional referral hospital and Benjamin Mkapa Hospital laboratory within three hours of sample collection, but haemolysed blood will not be processed; instead, venepuncture will be repeated. Next, the tourniquet will be removed, and the site of venepuncture will be pressed with a cotton swab to arrest bleeding. Centrifugation at 300 rpm for 5 minutes will be used to separate the serum sample from whole blood. Two aliquots will then be made, one for the lipid profile and the other for the serum electrolytes. A sample will be stored at 2^0^C – 8^0^C if the analysis is expected after 2 hours from sample collection, however, blood samples will be stored at room temperature if the analysis is expected to be done within 2 hours from sample collection. Using the clinical chemistry automated analyser, the sample will be analysed. German-made Elba machine XL-180 with serial number 160239. A high total cholesterol level of 200 mg/dL or higher, a low-density lipoprotein cholesterol level of 130 mg/dL or higher, a triglyceride level of 150 mg/dL or higher, or a high-density lipoprotein cholesterol level of 40 mg/dL for women and 50 mg/dL for males were all considered to be signs of dyslipidaemia [18].

The candidate’s palm is positioned palm-side up, the index, ring, or middle fingertip is chosen, and pressure is then applied to the fingertip to stimulate blood flow. After that, a fingertip is cleansed with methyl alcohol before a blood sample is drawn to measure blood sugar. This process is detailed in the sample collection manual SM-1-03.3 of the laboratory at Benjamin Mkapa Hospital and Dodoma Regional Referral Hospital. The finger will be held below the level of the elbow, pricked with a brand-new, sterile lancet to improve blood flow, and the gadget (ACCU-CHECK Active Roche glucometer machine) will be used to collect the blood directly from the puncture site. After the sample has been obtained, the customer will be given a ball of cotton wool to press on the finger for 10 minutes to stop the bleeding, and the lancet will then be disposed of in a sharp’s disposal box. Hyperglycaemia will be defined according to American Diabetes Association for non-diabetic patients hyperglycaemia will be defined as random blood sugar >11.1 mmol/L, or fasting blood sugar > 7.0 mmol/L and diagnosis of diabetes will be made with a fasting blood sugar ≥ 7.0 mmol/L, or random blood glucose ≥ 11.1 mmo/L plus symptoms of hyperglycaemia or glycated haemoglobin ≥ 6.5 %.

When processing samples, laboratory technicians at the BMH and the DRRH run controls in each machine daily to ensure the validity and reliability of the results. Essential maintenance is performed once every six months, whereas once a week is reserved for machine maintenance.

### Electrocardiogram (ECG)

The investigator will carry out a 12-lead ECG and is conversant with the manufacturer’s guidelines [17] under the direction of a professional cardiologist on each participant. Before a 12-lead ECG is taken, the patient will be told about the procedure, their privacy will be protected, and the environment will be kept comfortable to help the patient feel at ease and prevent interference with the ECG trace’s clarity. The necessary tools, such as the electrocardiograph, ECG paper, and ECG tabs, will be available to attach the electrodes and leads to the patient. The ECG cables must be kept from being twisted in order to prevent interference with ECG tracing. The patient will be directed to lie down at an angle of 45degrees with his or her head properly supported and the bed’s backrest, with the inner aspect of the patient’s wrist close to but not touching the patient’s waist. This is done after entering the patient’s ID number into the device and getting consent. As long as wet gel electrodes are utilised, shaving the skin won’t be necessary. The limb electrodes will next be placed in the following manner: Right inner wrist is red, left inner wrist is yellow, right inner leg is black just above the ankle, and left inner leg is green just above the ankle. The chest leads will be organised as follows: V4 is in the fifth intercostal space, mid-clavicular line, V5 is in the anterior axillary line, and V6 is in the mid-axillary line, the same horizontal line as V4 and V6. V1 is immediately to the right of the sternum, V2 is immediately to the left of the sternum, V3 is halfway between V4 and V2, V4 is in the fifth intercostal space, mid-clavicular line, and V6 is in the mid-axillary line, the same horizontal line as V4 and V5. ECG cables should not lie to each other and tension should be avoided to decrease artefact and increase the accuracy and quality of ECG tracing. The calibration signal on the ECG machine should be kept at a paper speed of 25 millimetre/second and ECG size 1 millivolt/10-millimeter deflection. During the procedure, the patient will be asked to remain motionless and breathe normally; the ECG trace should be clear prior to recording. The 12-lead ECG trace will include the patient’s name, hospital identification number, date of birth, and the day and time the ECG was taken [20]. According to the American College of Cardiology’s management guidelines for atrial fibrillation patients, the absence of P waves and an irregular-irregular RR interval are diagnostic signs of the condition [21], [22].

The Dutch organisation for Cardiology (2021) advises that the application date, patient data, relevant medical use, and the existence of a pacemaker or implantable cardioverter-defibrillator must all be met before utilising a Holter ECG monitor. It is important to time and record each symptom that a patient experiences. Additionally, the patient will become aware of alterations during the day, including variations in sleep, rest periods, and physical activity. The Dutch organisation for Cardiology (2021) advises that the application date, patient data, relevant medical use, and the existence of a pacemaker or implantable cardioverter-defibrillator must all be met before utilising a Holter ECG monitor. It is important to time and record each symptom that a patient experiences. Also, the patient will become aware of alterations during the day, including variations in sleep, rest periods, and physical activity.[23].

### Echocardiography

Only certain patients with ischemic stroke and additional characteristics, such as evidence of cardiac disease on history, examination, or electrocardiogram (ECG), suspected cardiac source of embolism (for example, infarctions in multiple cerebral or systemic arterial territories), suspected aortic disease, or paradoxical embolism, as well as patients with no other options, will be advised to undergo transthoracic echocardiography (model Vivid TM T9 made by GE Healthcare, USA, 2018)[24]

Only certain patients with ischemic stroke and additional characteristics, such as evidence of cardiac disease on history, examination, or electrocardiogram (ECG), suspected cardiac source of embolism (for example, infarctions in multiple cerebral or systemic arterial territories), suspected aortic disease, or paradoxical embolism, as well as patients with no other options, will be advised to undergo transthoracic echocardiography (model Vivid TM T9 made by GE Healthcare, USA, 2018)[25].

### Brain imaging

To confirm the stroke diagnosis, every patient will have an acute CT scan by SIEMENS (SOMATOM Definition Flash), and the majority will also get a brain MRI scan by MAGNETUM SPECTRA A TIM +Dot System 3T as part of a standard diagnostic procedure. Within the first 14 days following a stroke, patients with stroke-like symptoms but negative haemorrhagic stroke CT scan and unknown ischemic stroke status will be recruited for a study-specific MRI brain scan. The 3D-T1, axial T2, 3D-FLAIR, DWI, and SWI sequences make up the MRI study protocol. Before brain imaging, all patients will have their renal function status checked to lower the risk of contrast-induced nephropathy [26].The stroke volume (hematoma/infarct volume) will be calculated using the ellipsoid technique A+B+C/2, where A stands for the largest diameter, B for the largest diameter perpendicular to A, and C for the product of slice thickness and number of slices. While the volume is stated in millilitres or centimetres three, the lengths of A, B, and C are given in centimetres. [27], [28].

Bilateral regions of patchy or diffuse hypodensity on a CT scan or white matter hyperintensity on an MRI will be used to define Leukoaraiosis [29]. The global brain will be classified as absent if the third ventricle’s width is less than 5 mm, mild if it is between 5 and 6 mm, moderate if it is between 6 and 7 mm, and severe if it is greater than 7 mm [30]. All images will be downloaded to a computer workstation with a SYNGOVIA viewer, where two radiologists with the necessary training will review them.

### Study variables and measures

**Aim 1 Study Variables:** The variables address the prevalence of post-stroke depression at one month. MINI will be used for diagnosis of major depressive disorder (MDD) while PHQ-9 will be used to screen and measure the severity of depressive score (See Table 1 for a list of the variables with a description of aim 1)

**Aim 2 Study Variables:** The variables address the predictors of post-stroke depression at one month; these include age (in years), sex, alcohol use, cigarette smoking history, history of diabetes mellitus, dyslipidaemia, atrial fibrillation, post-stroke cognitive impairment and apathy, quality of life, stroke type and characteristic (haemorrhagic/ischaemic, cortical/sub-cortical), stroke (infarct/hematoma) volume, presence of Leukoaraiosis or brain atrophy (See table 2 for a list of the variables with a concise description of Aim 2)

**Aim 3 Study Variables:** The variables address the outcome of post-stroke depressive symptoms at three months, categorised as either improvement, significant worsening, or without significant change (See Table 3 for a list of the variables with a concise explanation for Aim 3).

**Aim 4 Study Variables:** The variables address the predictors of significant improvement of depressive symptoms at three months (See Table 4 for a list of the variables with a concise explanation for Aim 4).

### Dependent variable

Post-stroke depression:

### Primary dependent variable

Post-stroke depression will be defined as per PHQ-9 criteria; those with scores of ≥ 15 will be categorised as having major depressive disorder. Both severity and progression of depressive symptoms can be assessed with PHQ-9 tool [31].

### Secondary dependent variable

The outcome of PSD at three months will be evaluated by change of the PHQ-9 scores, categorised as a significant improvement if scores decrease by at least 5 points, significant worsening if scores increase by at least 5 points and no significant change if the scores remain within 5 points [32], [33].

### Assessment of change in Depressive symptoms

Patient Health Questionnaire (PHQ -9) will be used to assess the progress of depressive symptoms from baseline and after the three months of follow-up. The tool has a total score of 27, using the following nine items: little interest or pleasure in doing things, feeling down, depressed, or hopeless, trouble falling or staying asleep, or sleeping too much, feeling tired or having little energy, poor appetite or overeating, feeling bad about yourself or that you are a failure or have let yourself or your family down, trouble concentrating on things, such as reading the newspaper or watching television, moving or speaking so slowly that other people could have noticed and thoughts that you would be better off dead or of hurting yourself. Each item can score from 0-3, 0 if the participant replies (not at all), 1 if the participant responds (several days), 2 if responds (more than half the days), and 3 if participant responds (nearly every day). A final total score from each item is categorised as follows: a score of 1 -4 will be regarded as minimal depression, 5 – 9 as mild depression, 10 – 14 as moderate depression, 15 – 19 as moderately severe depression, and 20 – 27 as severe depression. PHQ-9 has been validated in Tanzania with a Sensitivity of 78% and a Specificity of 87%[34].

### Independent variables

Age (in years), sex, marital status, occupation, level of education, alcohol use, cigarette smoking history, diabetes mellitus, hypertension, dyslipidaemia, atrial fibrillation, type of stroke, lesion location, severity of stroke, stroke (infarct/hematoma) volume, presence of Leukoaraiosis, post-stroke cognitive impairment and apathy and quality of life.

### Assessment of Neurocognitive Functioning

The cognitive impairment will also be assessed using the Montreal Cognitive Assessment (MoCA), which is used to evaluate cognitive impairment, with the following domains: visuospatial/executive function(score of 5), naming(score of 3), attention(score of 6), language(repeat(score of 2) and fluency(score of 1), abstraction(score of 2), delayed recall(score of 5) and orientation(score of 6). Where by the optimal cut-off score point will be at a score of 22 with a sensitivity of 80% and specificity of 74%, and dementia at a score of 16, giving a sensitivity of 90% and specificity of 80% according to the validation of the tool done in Tanzania of which they used MoCA-5-min [35].

### Assessment of Apathy

Apathy will also be assessed using an evaluation scale which provides for behavioural, emotional and cognitive aspects of apathy. The tool comprises 18 items with a cut-off score of 39–41 with a sensitivity 75% and a specificity of 76.2% [36].

### Assessment of quality of life

Lawton-Brody Institutional Activities of Daily Living scale will be used to assess the patient capacity to perform tasks [37]. This tool includes almost eight domains of function like the ability to use the telephone, shopping, food preparation, laundry, mode of transportation, responsibility for own medications, housekeeping and ability to handle finances. Women will be scored on all eight areas of function. In contrast, for men, the areas of food preparation, housekeeping, and laundering will be excluded [38], with a sensitivity of 89% and specificity of 81% [39].

## Data Analysis

All data collected will be coded and entered into the computer for analysis. Data will be analysed using the Statistical Package for Social Sciences (SPSS) version 25.0. The data will be described by frequencies, proportions, Mean (± SD) and Median (IQR). The Chi-square test will be used to compare the association between post-stroke depression and the independent categorical predictors. To determine association between the independent variables and the post-stroke depression at one month and the outcome of depressive symptoms at three months, binary logistic regression analysis will be used, of which those variables with p < 0.2 under univariable analysis will be considered for multivariable analysis. Paired t-tests will be used to compare the mean changes in depressive symptoms from baseline to three months. Also, an odds ratio (OR) with a confidence interval of 95%, set at a significance level of < 0.05, will represent the results.

## Ethical approval and data dissemination

The ethical clearance was obtained from the institutional Research review committee of The University of Dodoma with the reference number MA.84/261/02 of 30/09/2022. Permission to conduct the study was provided by the administration of Dodoma Regional Referral Hospital (PB.22/1307/02/114) and Benjamin Mkapa Hospital (AB/150/293/01/391). All study participants will be required to sign written informed consent forms or proxy consent from a close relative or custodian in case the patient is incapable, which will state clearly about the conducted study. For confidentiality, participant names will not be utilised; only numbers will be used. Patients with the need for psychiatric including those with post-stroke depression will be referred for further evaluation and management will.

## Study timeline

The study will be conducted for 18 months from March 2025 to July 2026. Data collection will be for one year and three months, and follow-up for three months.

## Discussion

The prospective observation longitudinal nature of the study offers a robust temporal association of the prevalence, progression, and associated factors of post-stroke depression.

PHQ-9 screening tool is highly sensitive and specific in assessing the severity of depressive symptoms and their progression.

Given the nature of the study design and patients, the study has a risk of attrition, while has a strong capacity to elucidate the causal relationship between dependent and outcome variables. The study is time-consuming and expensive requiring close follow-up for accurate data collection.

The Dodoma University of Dodoma library, the study sites (Dodoma Regional Referral Hospital and Benjamin Mkapa Hospital), and a paper ready for submission in several peer-reviewed journals before publication will all receive the complete findings before they are published.

## Authors’ contributions

**Conceptualisation:** S.M, A.N

**Data curation:** S.M

**Formal analysis:** S.M

**Investigation:** A.N

**Methodology:** S.M, A.N

**Supervision**: A.N

**Writing – original draft:** S.M

**Writing – review, supervision and editing:** A.N

## Data Availability

Deidentified research data will be made publicly available when the study is completed and published.

## Acknowledgements

We would like to acknowledge the staff of Benjamin Mkapa Hospital and Dodoma Regional referral Hospital for all the assistance and also Dr Alphonce Baraka for his contribution in this work.

## Supporting Information

### Appendix

**Table 1:** Aim 1 Variable.

| Variable | Method of measurement | Operational definition | Level of measurement |
| --- | --- | --- | --- |
| Post-stroke depression at 1 month | Patient assessment | PHQ9(1-27), were by MDD likelihood for those who will score $\geq 15$ | Dichotomous |
| Major depressive disorder | M.I.N.I tool/questionnaire | Yes or No | Dichotomous |

**Table 2;.** Aim 2 Variable.

| Variable | Method of | Operational | Level of |
| --- | --- | --- | --- |

|  | **measurement** | **Definition** | **Measurement** |
| --- | --- | --- | --- |
| Age (years) | Medical records/patient report | Age in years | Continuous |
| Sex | Medical record/patient report | Male versus female | Dichotomous |
| Alcohol use | Patient report, interview | Yes/No | Dichotomous |
| Smoker | Patient report, interview | Yes/No | Dichotomous |
| Former smoker | Patient report, interview | who abstained from smoking for more than 12 months (Yes/No) | Dichotomous |
| Current smoker | Patient report, interview | Smoking in the past 12 months (Yes/No) | Dichotomous |
| Hypertension in (mmHg) | Medical record/patient report/assessment | Previously receiving antihypertensive medication or when the patient was previously diagnosed with hypertension or detecting blood pressure of $\geq 140/90$ mm/Hg for two measurements | Dichotomous |
| Diabetes | Patient report/medical records/blood sample | Yes/No | Dichotomous |
| Hyperlipidaemia | Medical record/patient report/blood | Previous had history of | Continuous |
| | sample | hyperlipidaemia or using lipid-lowering medication or total cholesterol $\geq 200$ mg/dl, LDL cholesterol $\geq 100$ mg/dl, and HDL-cholesterol $< 40$ mg/dl for men or $< 50$ mg/dl for women, and/or serum triglyceride level $\geq 150$ mg/dl | |
| Atrial fibrillation | Medical records | The absence of P waves and irregular-irregular RR interval | Dichotomous |
| Stroke severity | Patient report/assessment | NIHSS scale (1- 42) | continuous |
| Post-stroke cognitive impairment | Assessment | MoCA $< 22$ | Continuous |
| Apathy | Assessment | AES $> 38$ | Continuous |
| Stroke characteristics<br>1. Type<br>2. Volume of infarct/hematoma | Medical records | IS: hypodensity on CT scan/hypointense on MRI<br>ICH: hyperdense on CT scan/hyperintense on MRI<br>Stroke volume: ellipsoid method $A+B+C/2$, | Dichotomous<br>Continuous |
| Leukoaraiosis | Medical records | Bilateral Areas of patchy or diffuse hypodensity on CT or white matter hyperintensity on | Dichotomous |
|  |  | MRI |  |
| Brain atrophy | Medical records | The width of the third ventricle is greater than 5 mm | Dichotomous |
| Type of medication for stroke received | Medical records/patient report | Name of the medication | Nominal |

**Table 3:** Aim 3 Variable.

| **Variable** | **Method of measurement** | **Operational definition** | **Level of measurement** |
| --- | --- | --- | --- |
| Change in depressive symptoms at 3 months post-stroke | Patient assessment | PHQ9(1-27), 5-point change will be considered as significant change | Continuous |

**Table 4:** Aim 4 Variable.

| **Variable** | **Method of measurement** | **Operational definition** | **Level of measurement** |
| --- | --- | --- | --- |
| Predictors of significant improvement of depressive symptoms at 3 months | Binary logistic regression | Which independent variable is statistically significant associated with significant improvement of depressive symptoms | Continuous and dichotomous |

### Appendices 2

#### Appendix 2: Care Report Form/ questionnaire

Name............................................................

Date of admission.......... /............. /...............

Hospital reg # ..............................................

Contacts

1..............................2.....................................3..............................

1. Demographic data

1. Patient’s Identification Number................................................

2. Patient’s residence: a) Urban b. Rural

3. Date of birth ............/............/............

4. Sex a) Male b. Female

5. Marital status a. Single b. Married c. divorced/separated d. widowed/ widower

6. Level of education a. No formal school b. Primary school c. Secondary school d.

Tertiary school

2. Risk factors & past medical history

7. Cigarette smoking a. Yes, b. No

8. If “yes” number of cigarettes per day..................

9. Number of years smoked ..............................

10. Pack years................

11. Alcohol consumption a. Yes, b. No

12. If “Yes” For how long? ..................(Years)

13. Units per day/week... 86

14. Hypertension a. Yes b. No

15. Diabetic Mellitus a. Yes b. No

3. Clinical presentation on admission

16. Headache

a. Yes b. No

17. Aphasia

a. Yes b. No

18. Nausea/vomiting

a. Yes b. No

19. Loss of consciousness

a. Yes, b. No

20. Focal neurological deficit/Limb weakness

a. Yes, b. No

21. Dysarthria

a. Yes b. No

22. Seizures/ convulsion

a. Yes, b. No

23. Pupil examinations

a. Normal b. Anisocoria c. Pinpoint

4. Investigations

CT scan results

24. Type of stroke a) haemorrhagic b) ischemic (If the type of stroke is haemorrhagic then respond to Qn No, 25 - 26)

25. Location of haemorrhagic stroke

a) Lobar

b) Non-lobar

26. Volume of hematoma __________ cm□/millilitre (if the type of stroke is ischemic respond to Qn No, 27 – 30

27. Infarct volume __________ cm□/millilitres Vital Signs 1. Heart Rate ____ bpm 2.

Pulse rate ____ bpm 3. Pulse deficit ____ bpm 4. BP (mmHg) ____/____ 5. Mental

status (GCS) ____/15 87

28. Involvement of strategic site (thalamus, angular gyrus, cingulate gyrus, caudate,

Globus pallidus, basal forebrain, anterior limb of the internal capsule, or hippocampus) (put a tick where appropriate) a) Yes b) No

29. Number of infarcts (give the actual number) ..................

30. If multiple, where are they located? a) dominant hemisphere b) nondominant hemisphere c) both hemispheres d) anterior circulation e) posterior circulation f)anterior and posterior circulation

31. Presence of Leukoaraiosis

a) Yes b) No

32. Presence of global brain atrophy

a) Yes b) No

33. ECG Results..................................................................

34. Random Blood Glucose (results) .....................mmol/l

35. Cholesterol................... mmol/l

36. LDL .....................mmol/l

37. HDL .....................mmol/l

38. Triglyceride.....................mmol/

39. History of aspiration pneumonia (tick where appropriate)

a. Yes b. No

40. Current medications the patient is on

a. .................................

b. .................................

c. ..................................

d. ..................................

e. ..................................

41. Treatment of stroke

a. .................................

b. .................................

c. .................................

d. .................................

e. .................................

#### Appendix 3 : National Institute of Health Stroke Scale NIHSS

Instructions Case definition Score

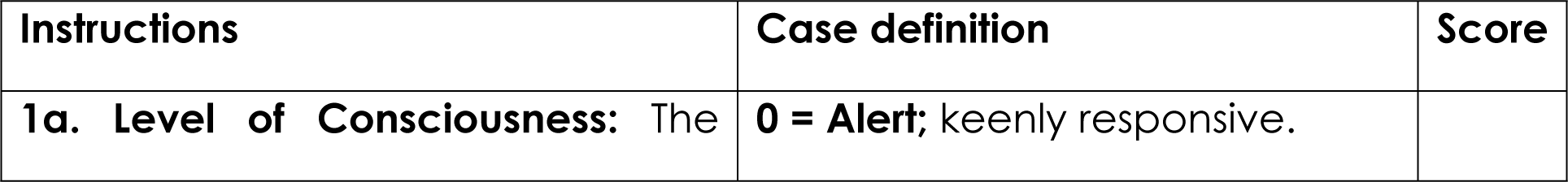
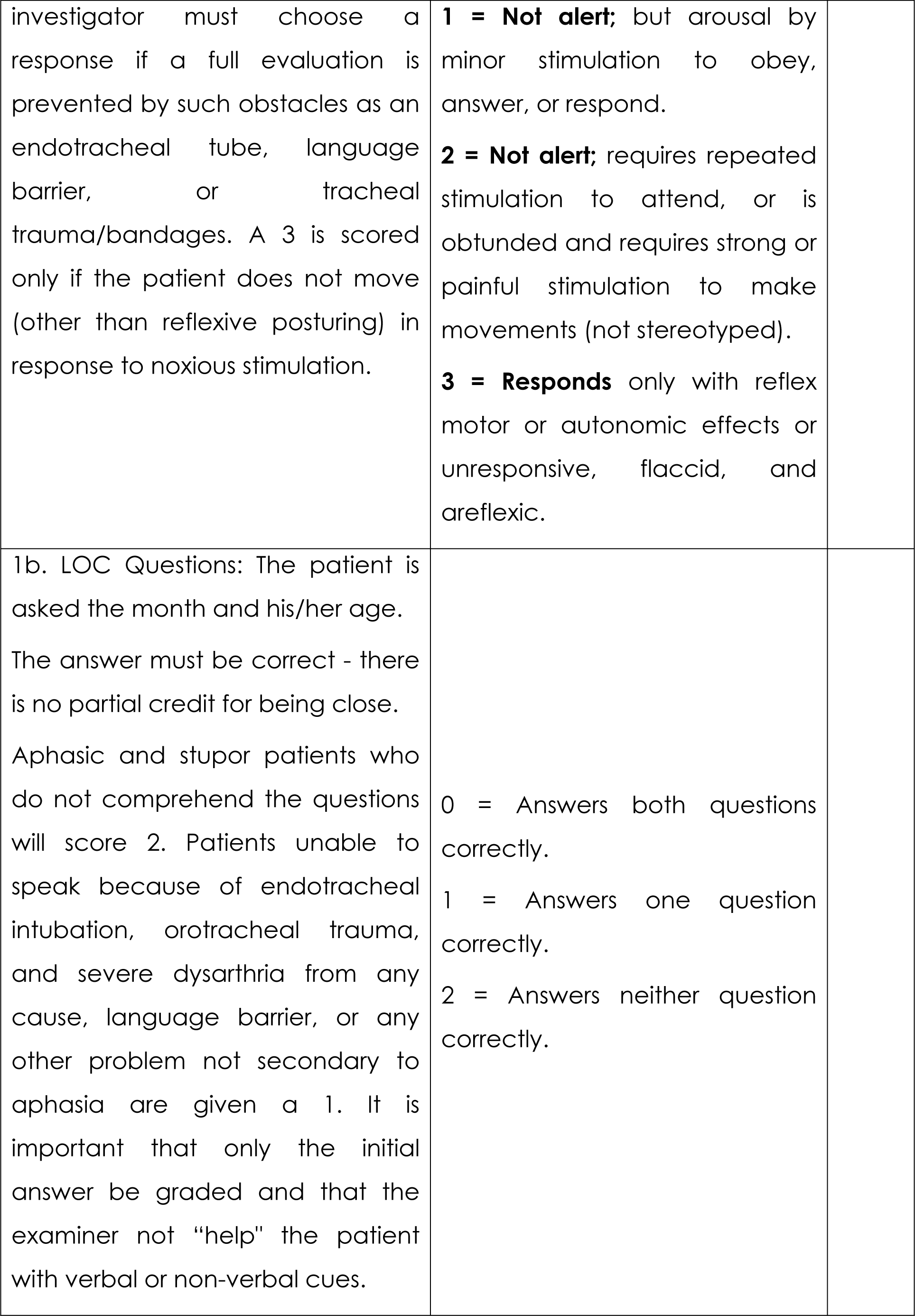
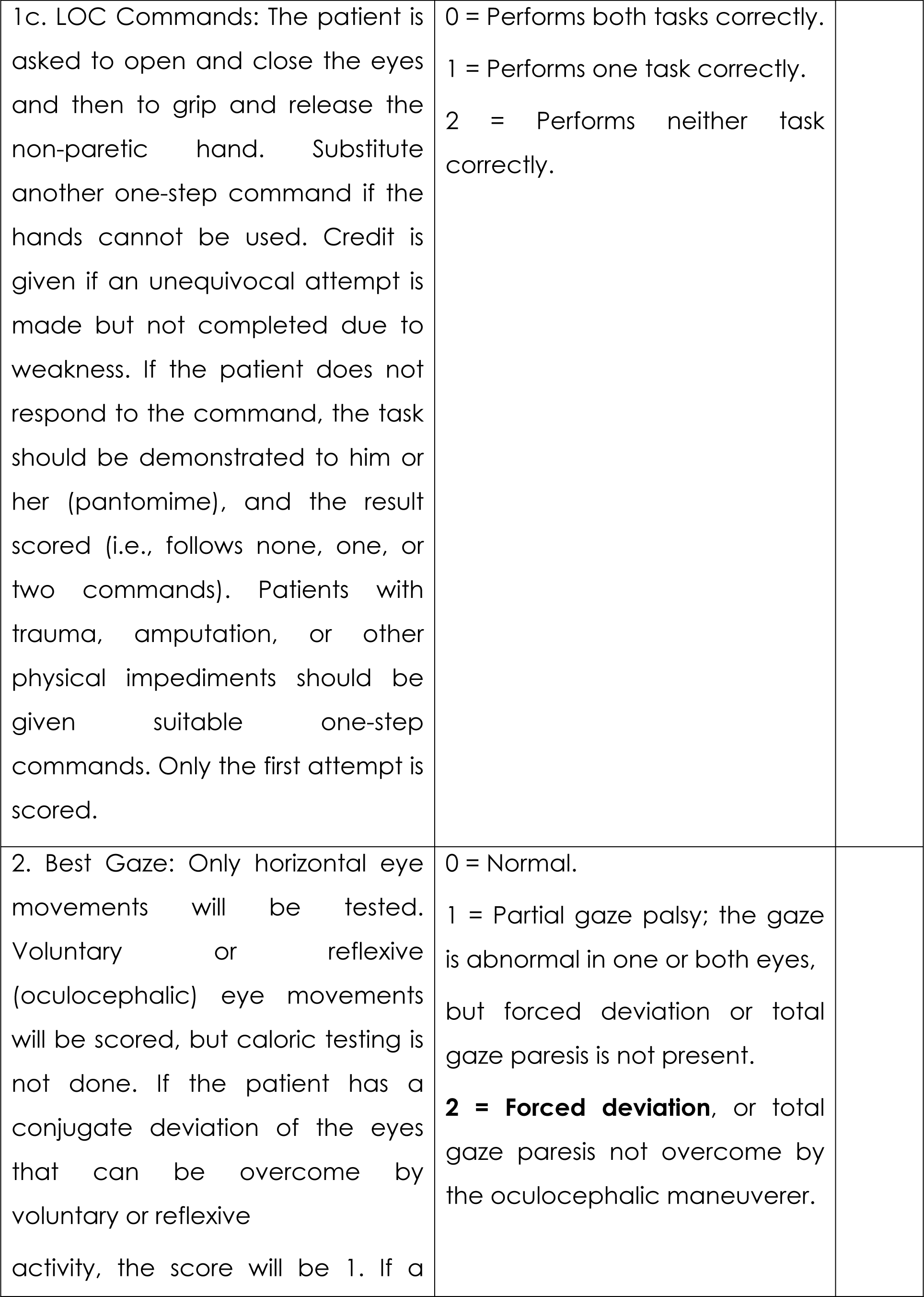
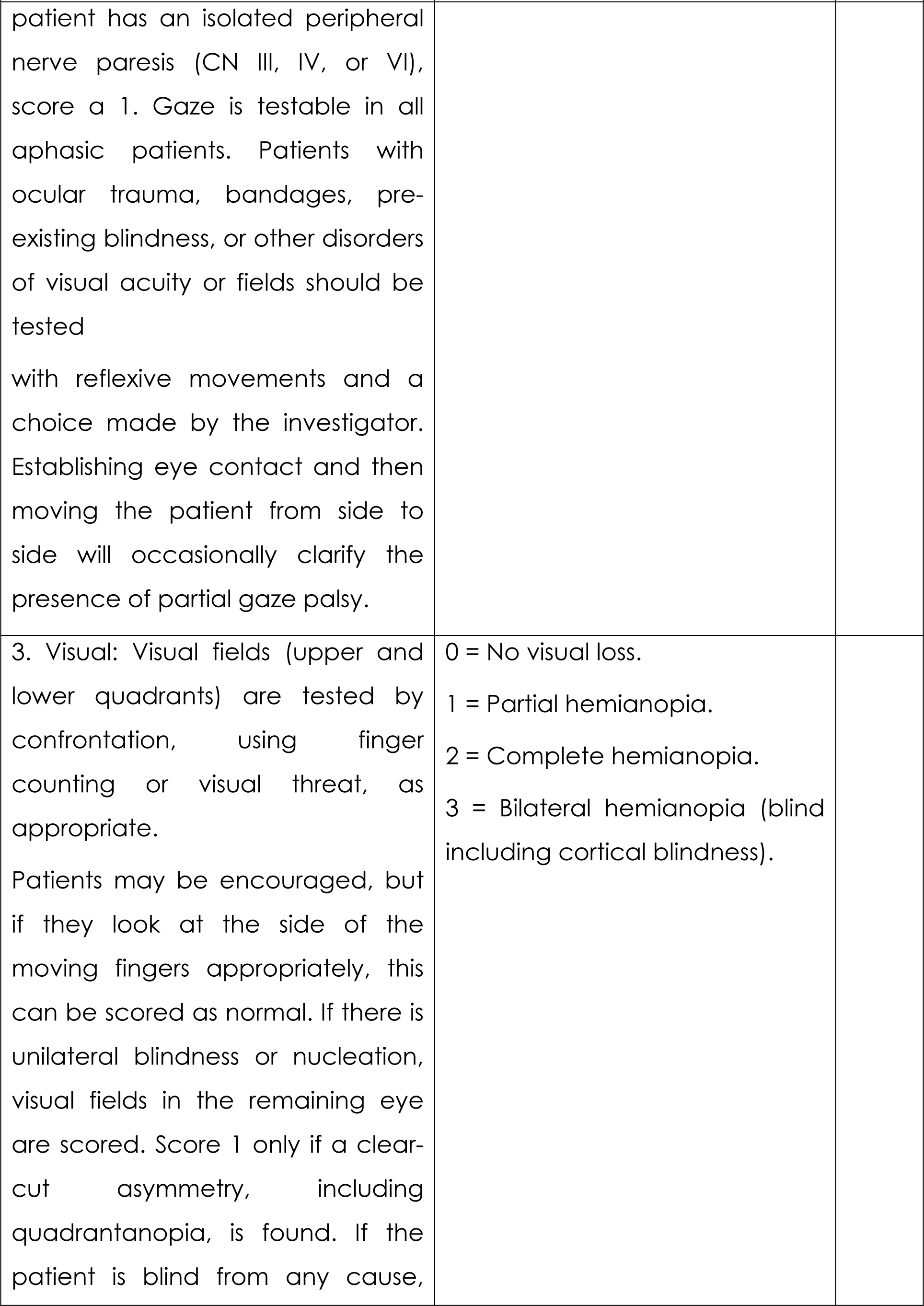
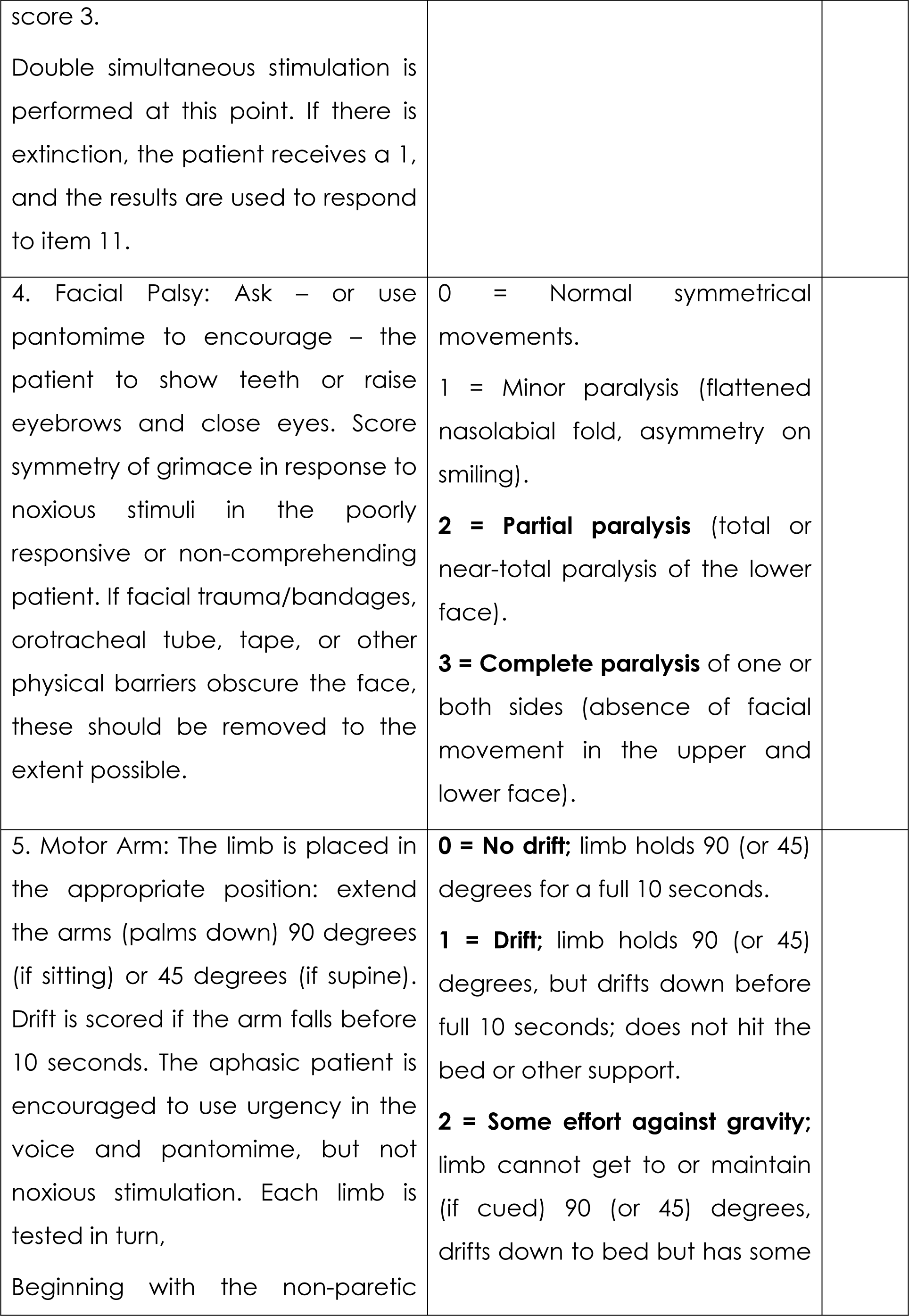
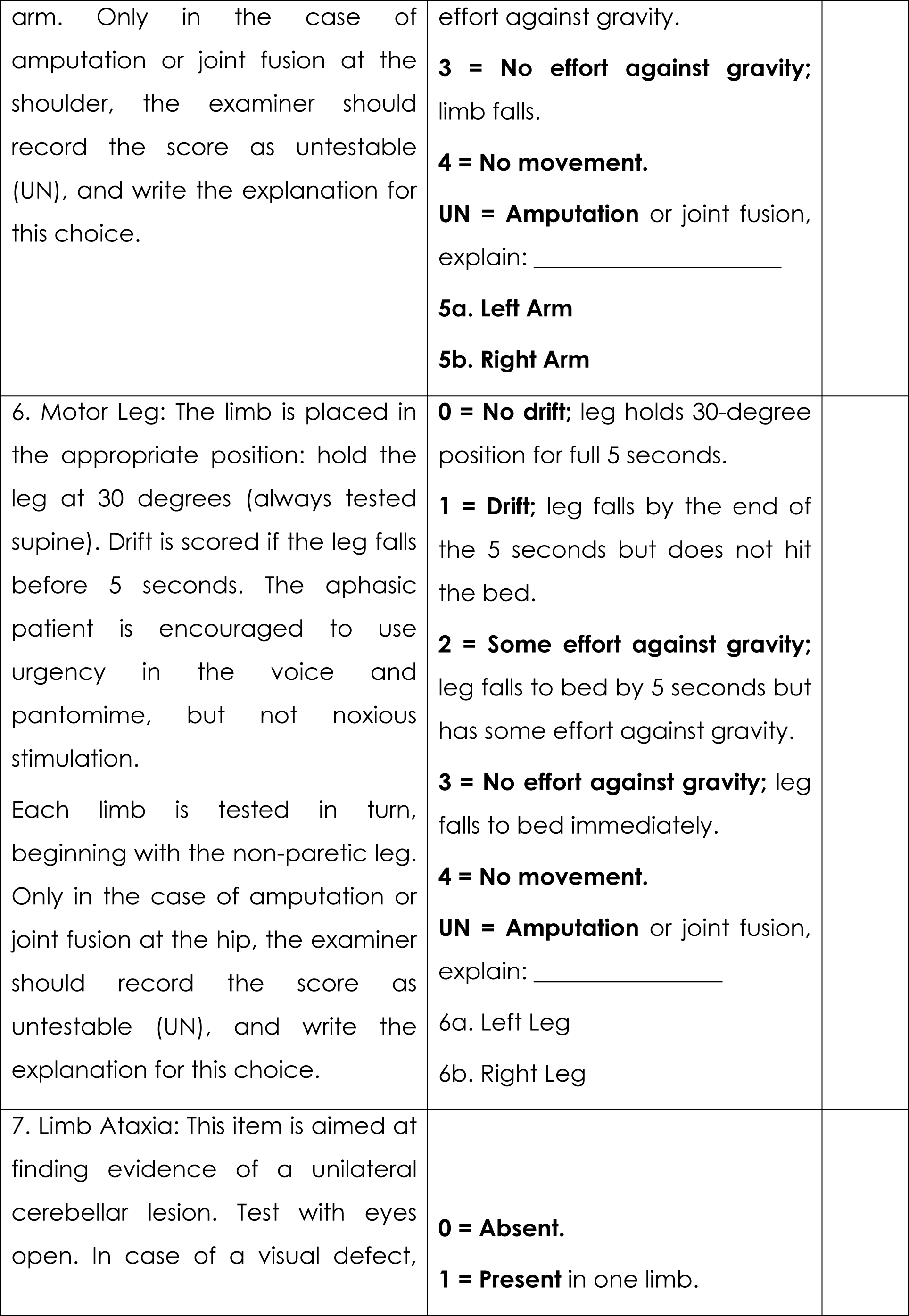
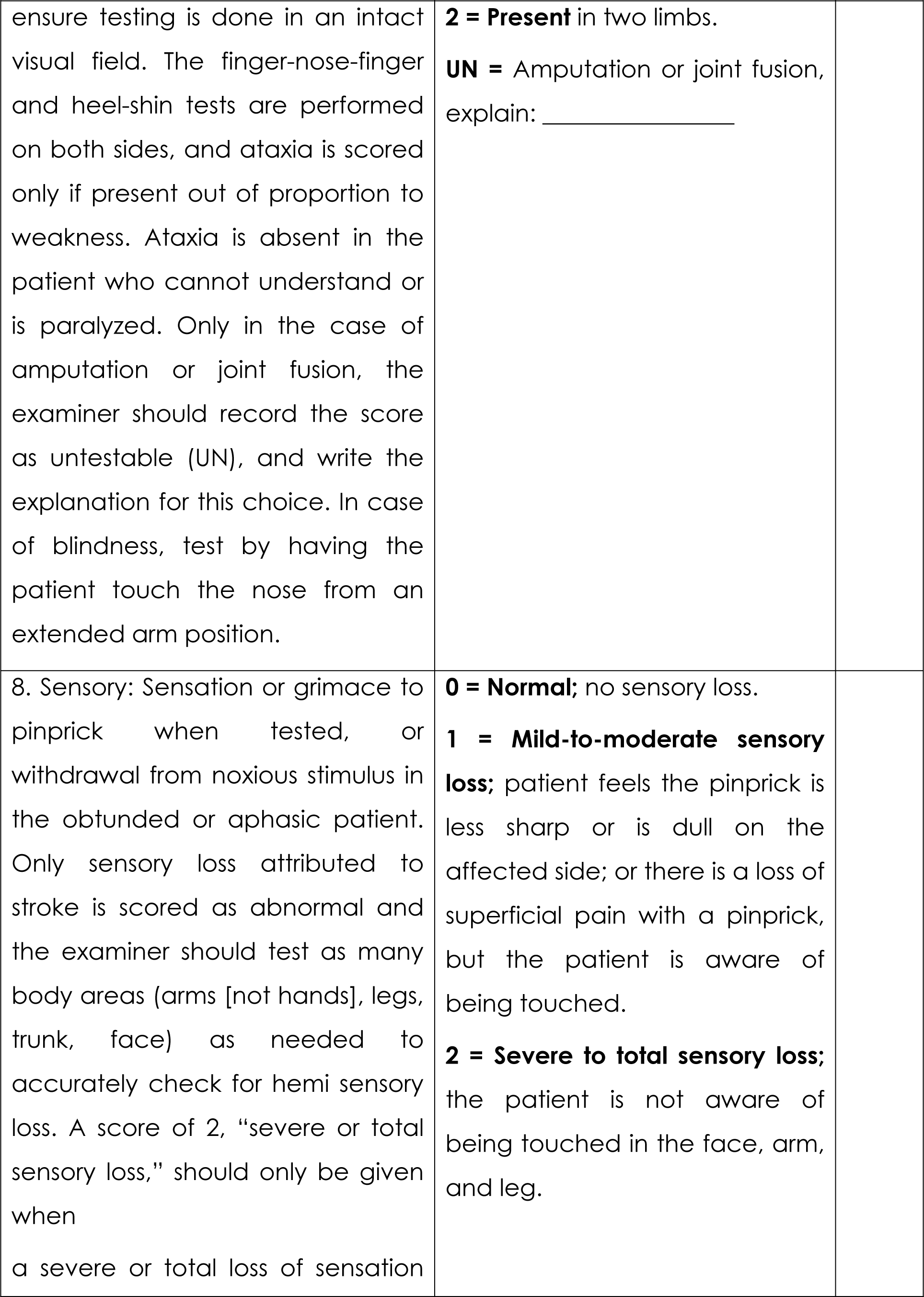
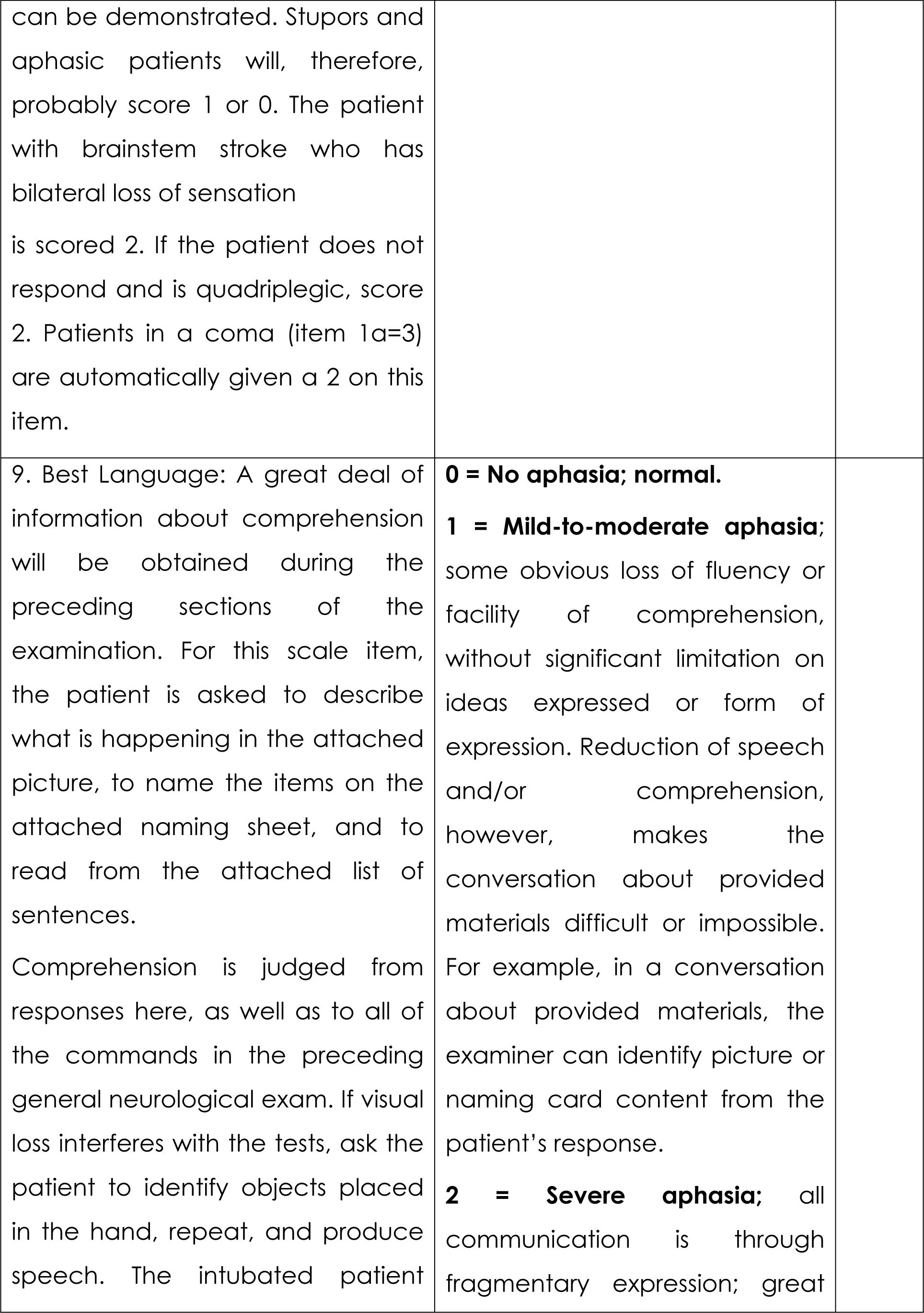
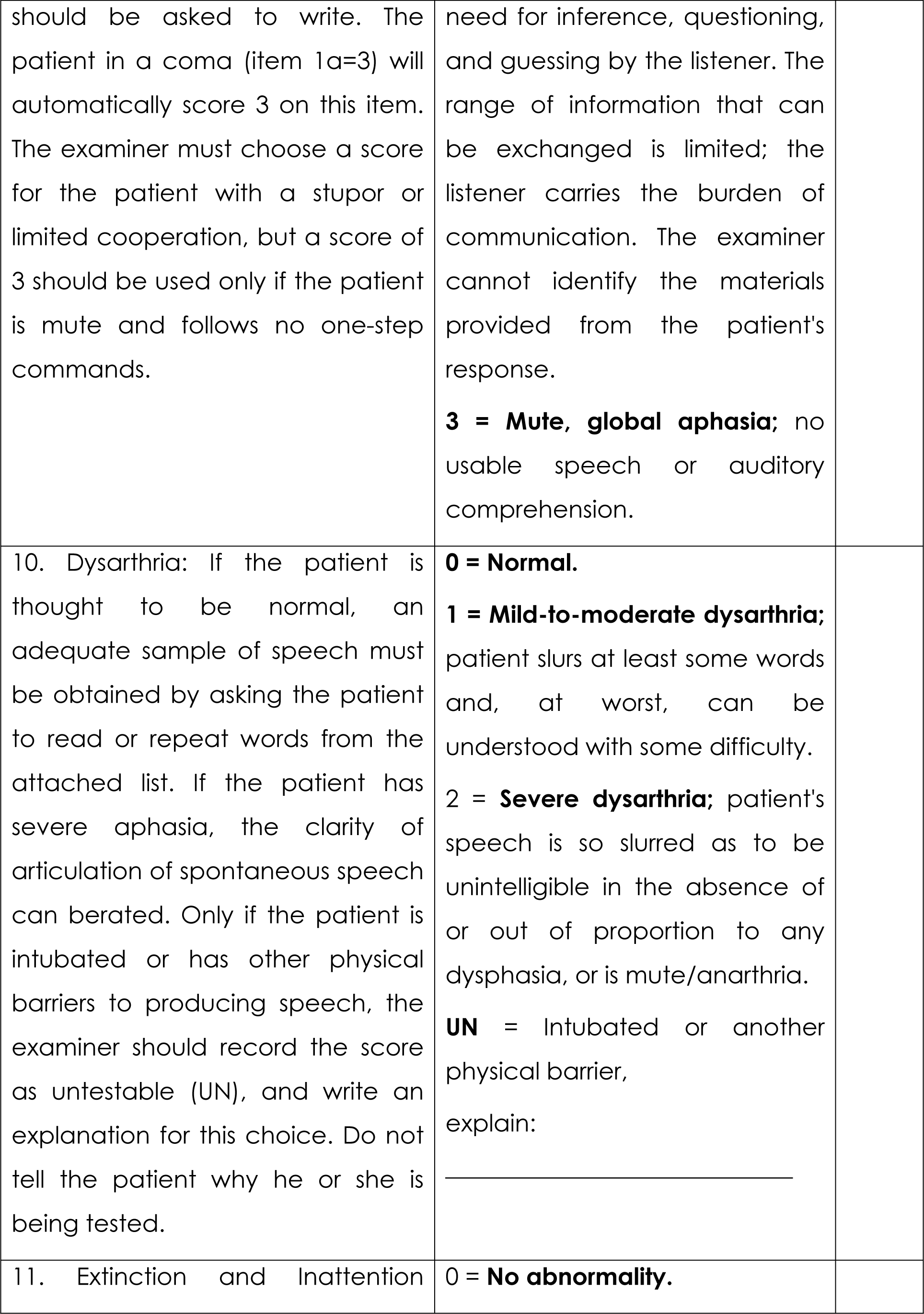
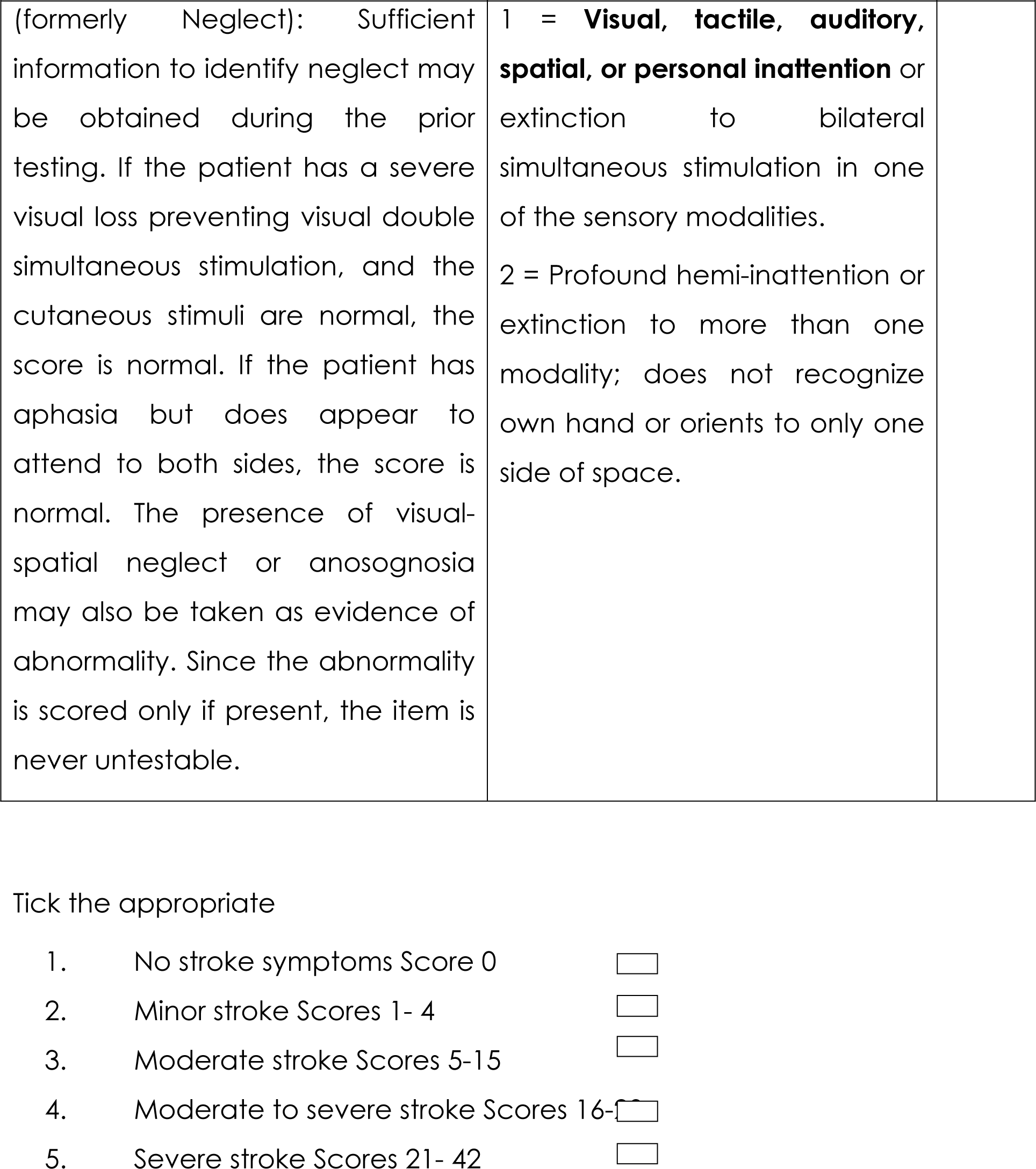

#### Appendix 4 : Apathy Evaluation Scale (AES)

Date ____________________ ID No ____________________

Rate each item based on an interview of the subject. The interview should begin with a description of the subject’s interests, activities, and daily routine. Base your ratings on both verbal and non-verbal information. Ratings should be based on the past 4 weeks. For each item ratings should be judged:

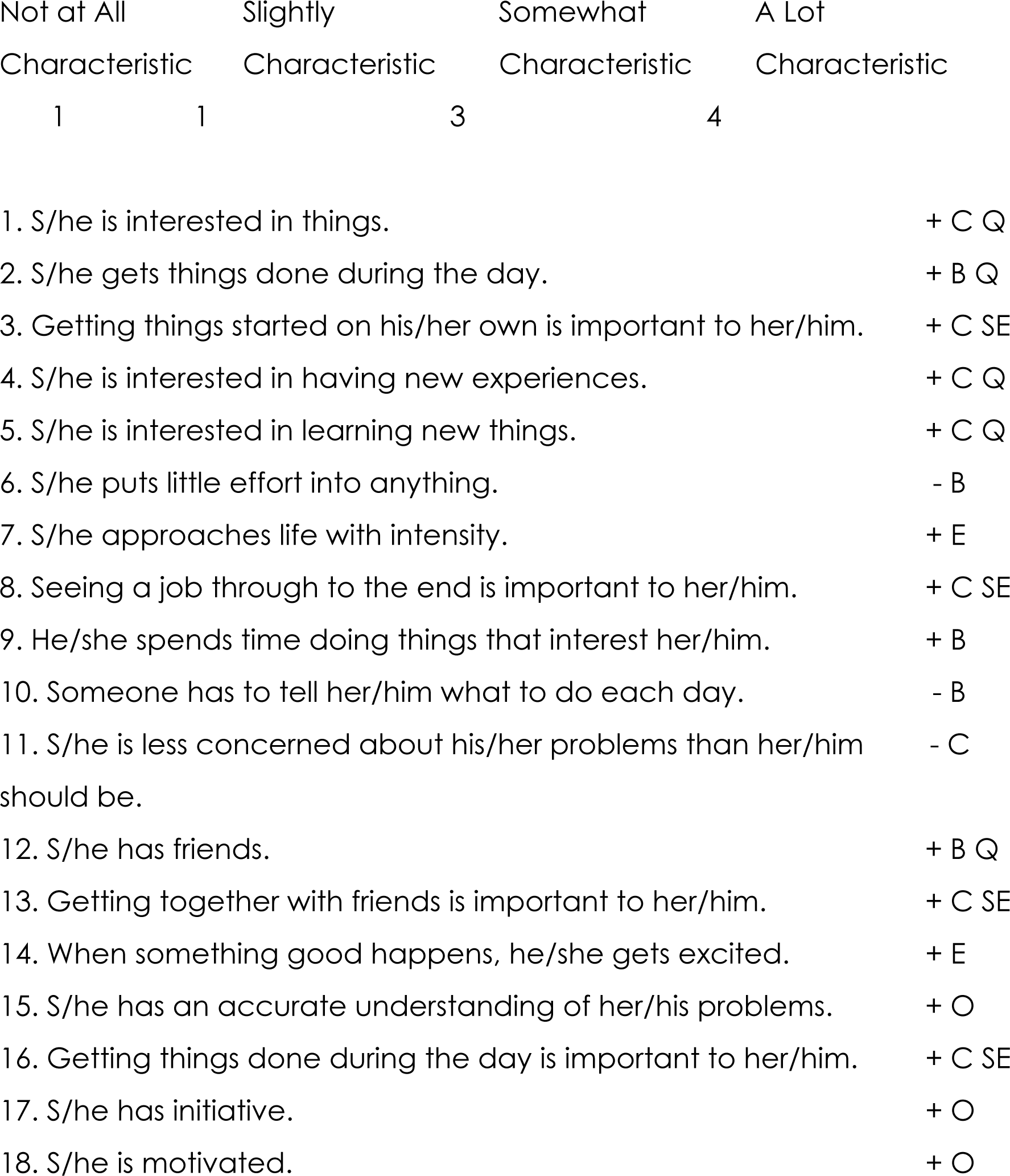
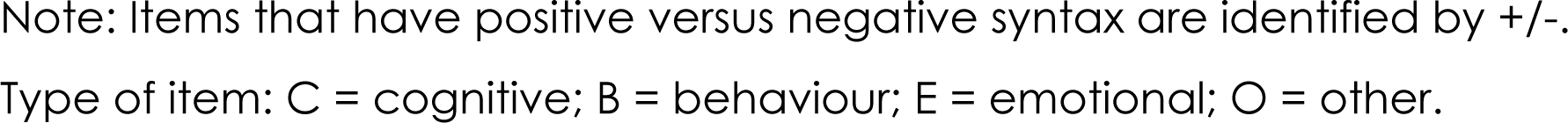

#### Appendix 5 : The Patient Health Questionnaire - 9

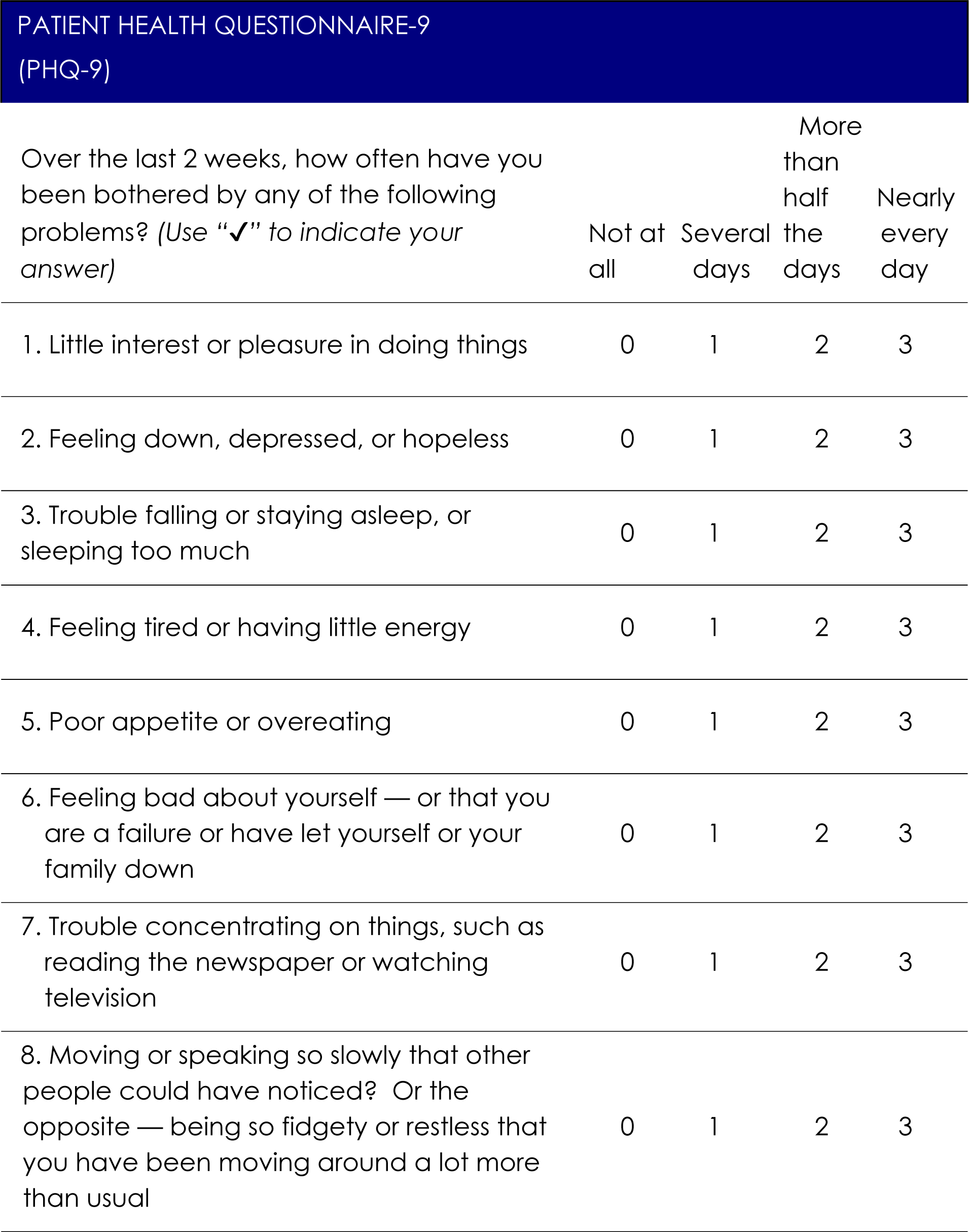
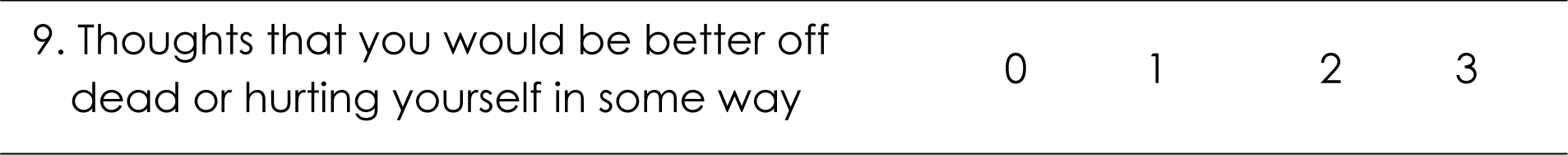

#### Appendix 6: Montreal Cognitive Assessment (MoCA) test

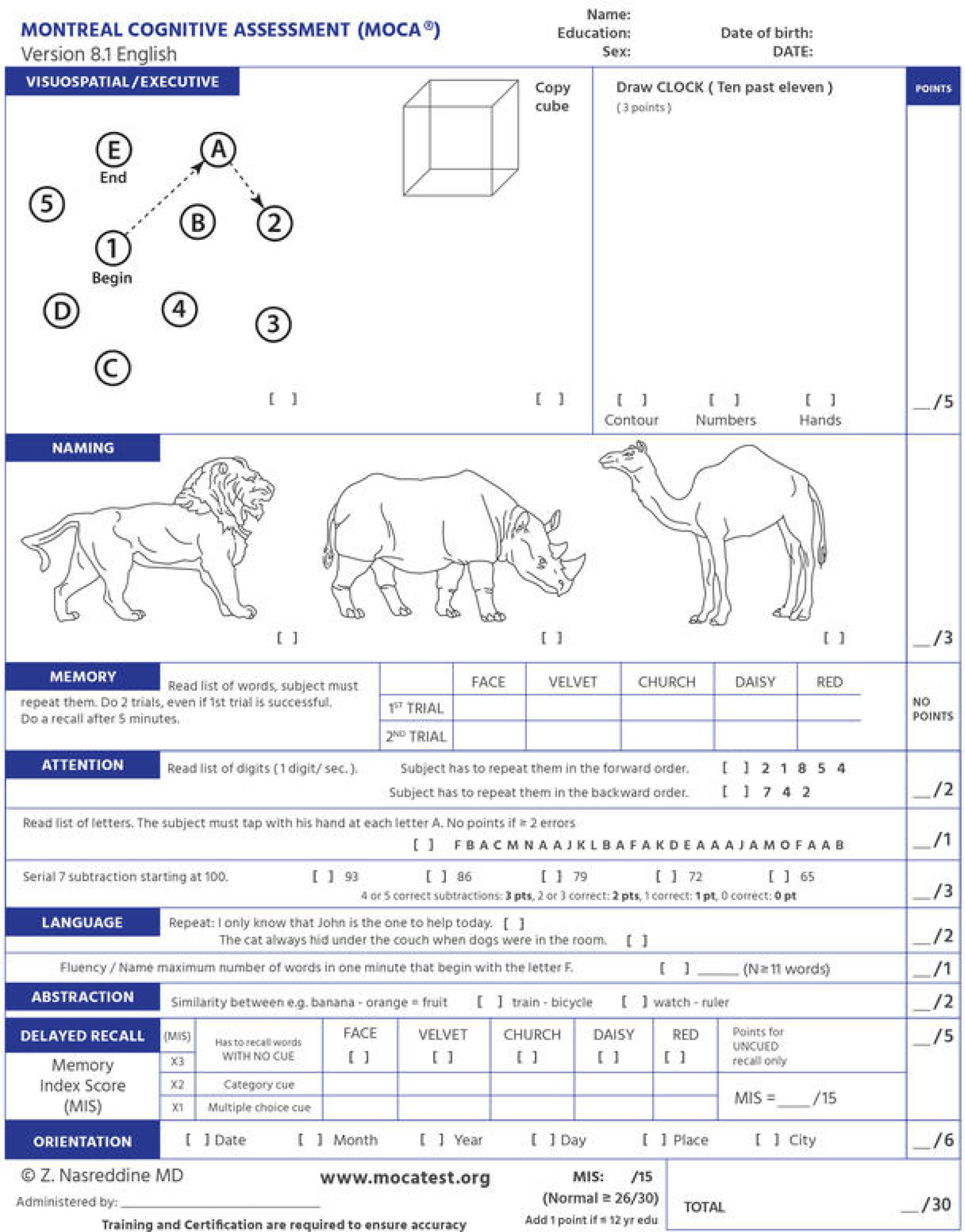

